# Antithrombotic Therapy After Heart Valve Surgery, Contemporary Practice in the United Kingdom

**DOI:** 10.1101/2022.12.05.22283122

**Authors:** Nabila Laskar, Christopher D Bayliss, Enoch Akowuah, Bilal H Kirmani, John B Chambers, Rebecca Maier, Norman P Briffa, Neil Cartwright, Simon Kendall, Benoy Nalin Shah

## Abstract

**Objectives:** There is a lack of high-quality data informing the optimal antithrombotic drug strategy following bioprosthetic heart valve replacement or valve repair. Disparity in recommendations from international guidelines reflects this. This study aimed to document current patterns of antithrombotic prescribing after heart valve surgery in the United Kingdom.

**Methods:** All UK consultant cardiac surgeons were e-mailed a custom-designed survey. The use of oral anticoagulant (OAC) and/or antiplatelet drugs following bioprosthetic aortic (AVR) or mitral valve replacement (MVR), or mitral valve repair (MVrep), for patients in sinus rhythm, without additional indications for antithrombotic medication, was assessed. Additionally, we evaluated anticoagulant choice following MVrep in patients with atrial fibrillation (AF).

**Results:** We identified 260 UK consultant cardiac surgeons from 36 units, of whom 103 (40%) responded, with 33 units (92%) having at least one respondent. The greatest consensus was for patients undergoing bioprosthetic AVR, in which 76% of surgeons favour initial antiplatelet therapy and 53% prescribe lifelong treatment. Only 8% recommend initial OAC. After bioprosthetic MVR, 48% of surgeons use an initial OAC strategy (versus 42% antiplatelet), with 66% subsequently prescribing lifelong antiplatelet therapy. After MVrep, recommendations were lifelong antiplatelet agent alone (34%) or following 3 months OAC (20%), no antithrombotic agent (20%), or 3 months OAC (16%). After MVrep for patients with established AF, surgeons recommend warfarin (38%), a direct oral anticoagulant (37%) or have no preference between the two (25%).

**Conclusion:** There is considerable variation in the use of antithrombotic drugs after heart valve surgery in the UK. This reflects the lack of high-quality evidence and underscores the need for randomised studies.

**KEY MESSAGES:** *What is already known on this topic?:* The most appropriate antithrombotic drug regime following bioprosthetic heart valve replacement or mitral valve repair is not known. Contemporary practice in the UK has not been established.

*What this study adds:* This study demonstrates wide variation in antithrombotic drug choice and duration following bioprosthetic heart valve replacement or mitral valve repair in the UK.

*How this study might affect research, practice, or policy:* Randomised studies are required to assess the most appropriate antithrombotic strategy after heart valve surgery.

## INTRODUCTION

The first two decades of this century have witnessed a shift in the choice of prosthetic heart valves amongst patients undergoing surgery for heart valve disease (HVD), with a fall in the use of mechanical valves and reciprocal increase in use of bioprosthetic valves and valve repair^1^. A prominent factor driving this change is the desire of patients to avoid a lifetime commitment to anticoagulation with warfarin.

Although it is widely accepted that anticoagulation with a vitamin K antagonist (VKA) is required for patients with mechanical heart valves to prevent thromboembolic complications, the development of thrombi on biological valves is also a recognised phenomenon. This risk is greatest in the initial period after surgery^2^, prior to endothelialisation of prosthetic material. Recent imaging studies have highlighted the hitherto under-recognised presence of subclinical leaflet thrombosis, often termed hypo-attenuated leaflet thickening (HALT), which can be seen on both surgical and transcatheter bioprostheses and may affect long-term valve durability^3^.

The choice of antithrombotic agent after bioprosthetic aortic (AVR) or mitral valve replacement (MVR), or mitral valve repair (MVrep) for patients in sinus rhythm is controversial, with only observational data and small prospective studies (often with conflicting outcomes) available to guide practice.^4-8^ The 2020 American (ACC/AHA) and 2021 European (ESC/EACTS) clinical guidelines on valvular heart disease differ in their recommendations. The American guidelines suggest antiplatelet therapy or a short course (up to 6 months) of oral anticoagulation (OAC) for both bioprosthetic AVR and MVR (provided there is low risk of bleeding and no other indication for OAC), with no recommendation for MVrep. European guidelines suggest a similar strategy for bioprosthetic AVR, albeit for a shorter duration (3 months), but recommend a vitamin K antagonist for the first 3 months following bioprosthetic MVR or MVrep.^9,10^

Anecdotal evidence suggests a substantial variation in practice. The aim of this study was therefore to provide objective evidence of current practice from a cross-sectional survey of UK cardiac surgeons.

## METHODS

The survey was designed by a team of cardiologists and cardiac surgeons. It comprised ten questions relating to current departmental policy and individual surgeon practice for antithrombotic drug use after bioprosthetic AVR, MVR and MVrep in patients in sinus rhythm with no other indication for antithrombotic medication. Additional questions included the choice of OAC (vitamin K antagonist versus direct oral anticoagulants) in patients undergoing mitral valve repair with pre-existing atrial fibrillation (AF).

Using the Society for Cardiothoracic Surgery’s (SCTS) directory, NHS Trust websites and telephone contacts, all consultant cardiac surgeons in the UK who perform heart valve operations on adult patients were contacted. The survey (see supplemental material) was compiled using online software (Google LLC, California, USA) and a link e-mailed to all surgeons. One month later, following fortnightly reminder emails, responses were collated and analysed. As this study did not involve the use of any patient data, ethical approval was not required.

## RESULTS

From the 36 surgical centres in the UK, we identified 260 consultant cardiac surgeons (94% male) performing heart valve surgery on adult patients, of whom 103 (40%) completed the survey. No replies were received from any consultant in 3 centres.

Surgeons from 10/33 centres confirmed they have a departmental policy for antithrombotic drug prescribing after heart valve surgery. There was no such policy in 11/33 centres and there was disagreement as to whether policy existed or not in the remaining 12 centres. We found that most centres (23/33, 70%) had variation between their individual surgeons, both in choice of agent and duration of therapy.

Initial strategies can be seen in Figure 1 and Figure 2, with a full breakdown of antithrombotic regimes utilised demonstrated in Table 1. The greatest consensus was seen amongst patients undergoing bioprosthetic AVR, for whom 76% of surgeons favour initial antiplatelet therapy, with 53% recommending lifelong treatment. No antithrombotic agent is favoured by 17 surgeons (17%) whilst 8% of surgeons recommended 3 months of OAC followed by lifelong antiplatelet treatment. The choice of OAC (vitamin K antagonist or novel oral anticoagulant), if used, is demonstrated in Table 2.

**Table 1.**
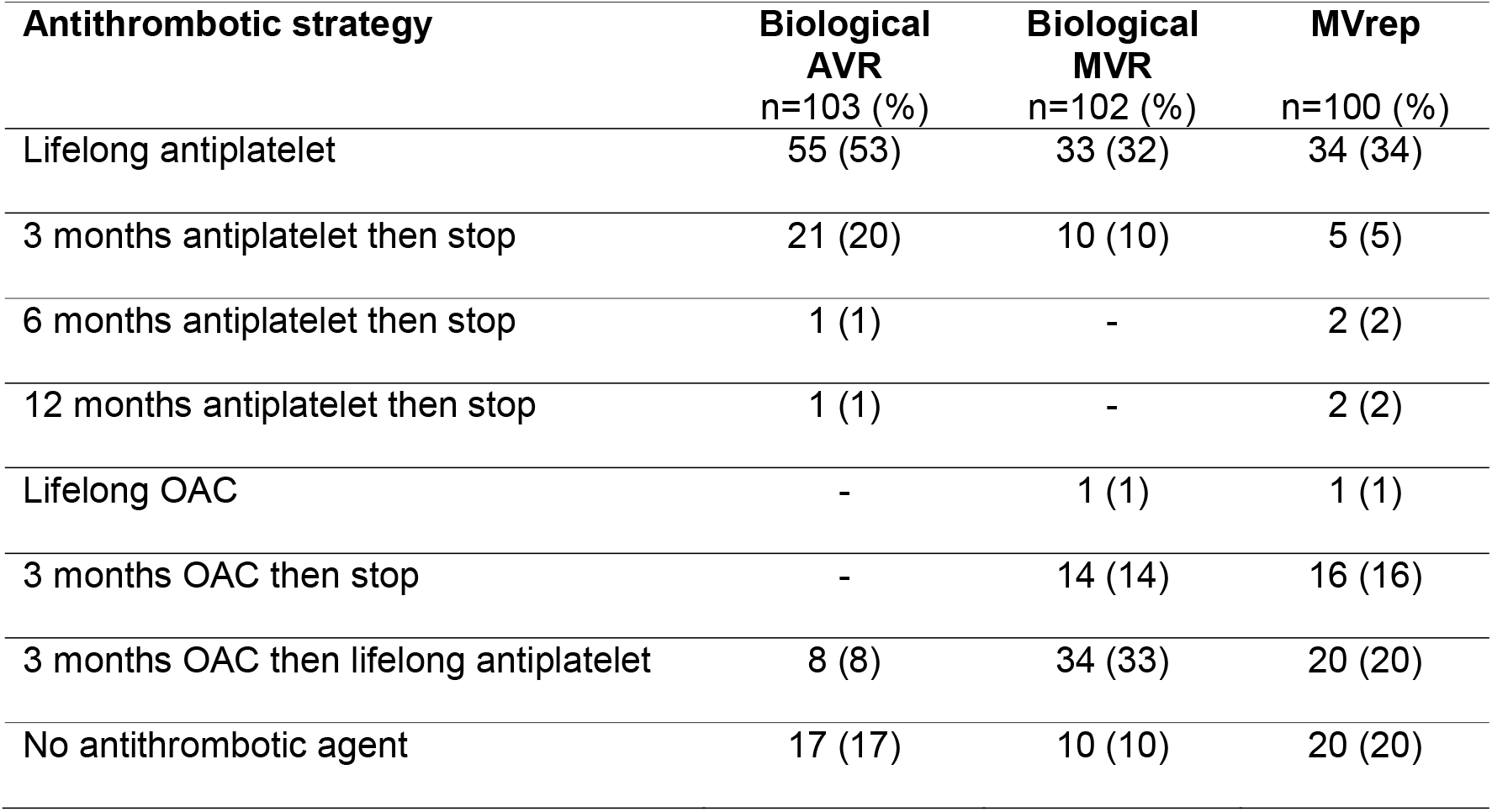
Choice of antithrombotic agent following bioprosthetic heart valve surgery or mitral valve repair in patients without another indication for anticoagulation. AVR, aortic valve replacement; MVR, mitral valve replacement; MVrep, mitral valve repair; OAC, oral anticoagulant.

**Table 2.** Choice of oral anticoagulant following bioprosthetic heart valve surgery or mitral valve repair in patients without another indication for anticoagulation. AVR, aortic valve replacement; MVR, mitral valve replacement; MVrep, mitral valve repair; DOAC, direct oral anticoagulant.

| **Oral anticoagulant strategy** | **Biological**<br>**AVR**<br>**n=8 (%)** | **Biological**<br>**MVR**<br>**n=49 (%)** | **MVrep**<br>**n=34 (%)** |
| --- | --- | --- | --- |
| Warfarin | 4 (50) | 34 (69) | 24 (71) |
| DOAC | 2 (25) | 8 (16) | 7 (21) |
| Either (no preference) | 2 (25) | 7 (14) | 3 (9) |

**Figure 1.**
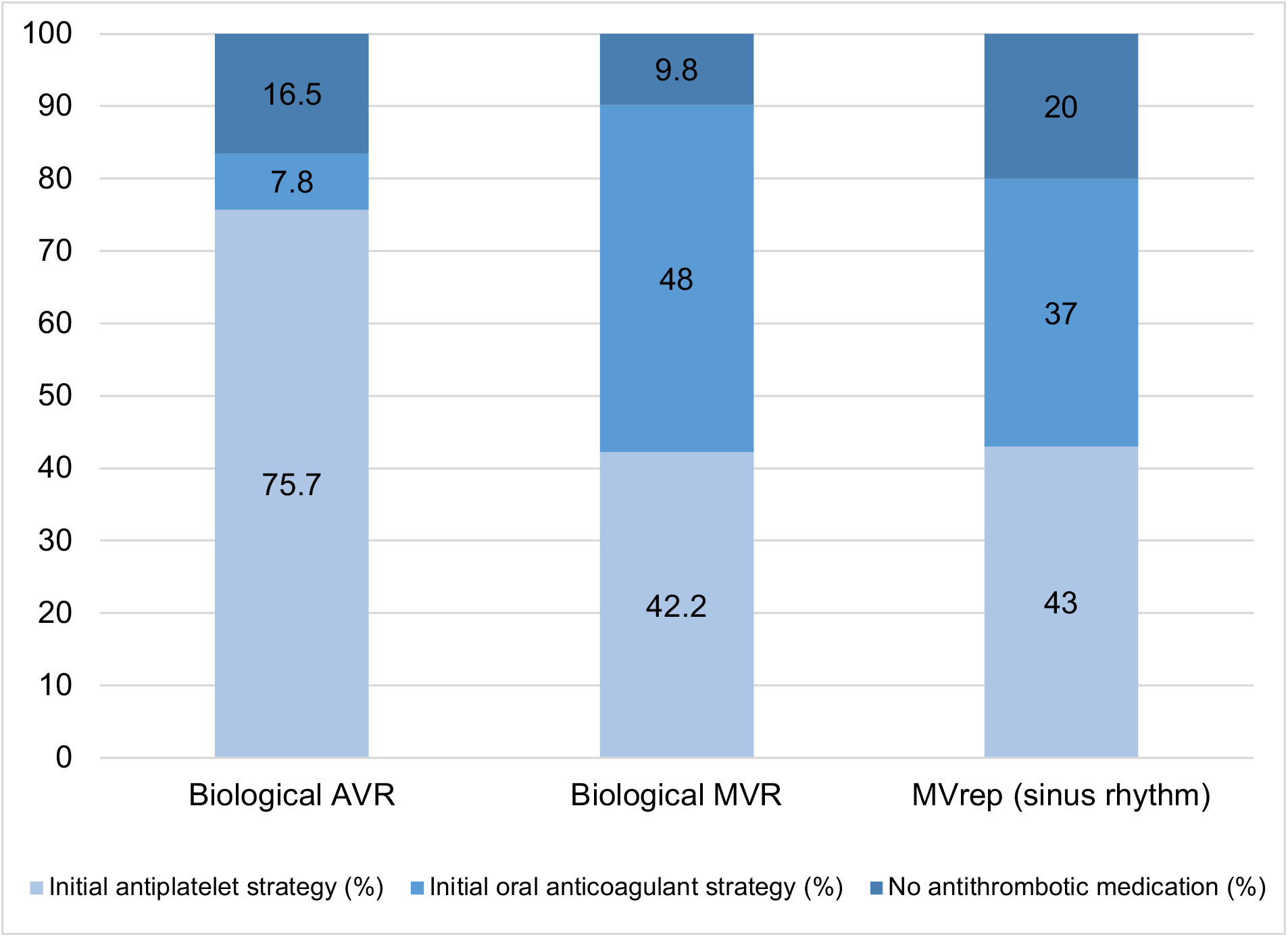
Summary graph showing initial antithrombotic strategy following bioprosthetic heart valve surgery or mitral valve repair in patients without another indication for anticoagulation. AVR, aortic valve replacement; MVR, mitral valve replacement; MVrep, mitral valve repair.

**Figure 2.**
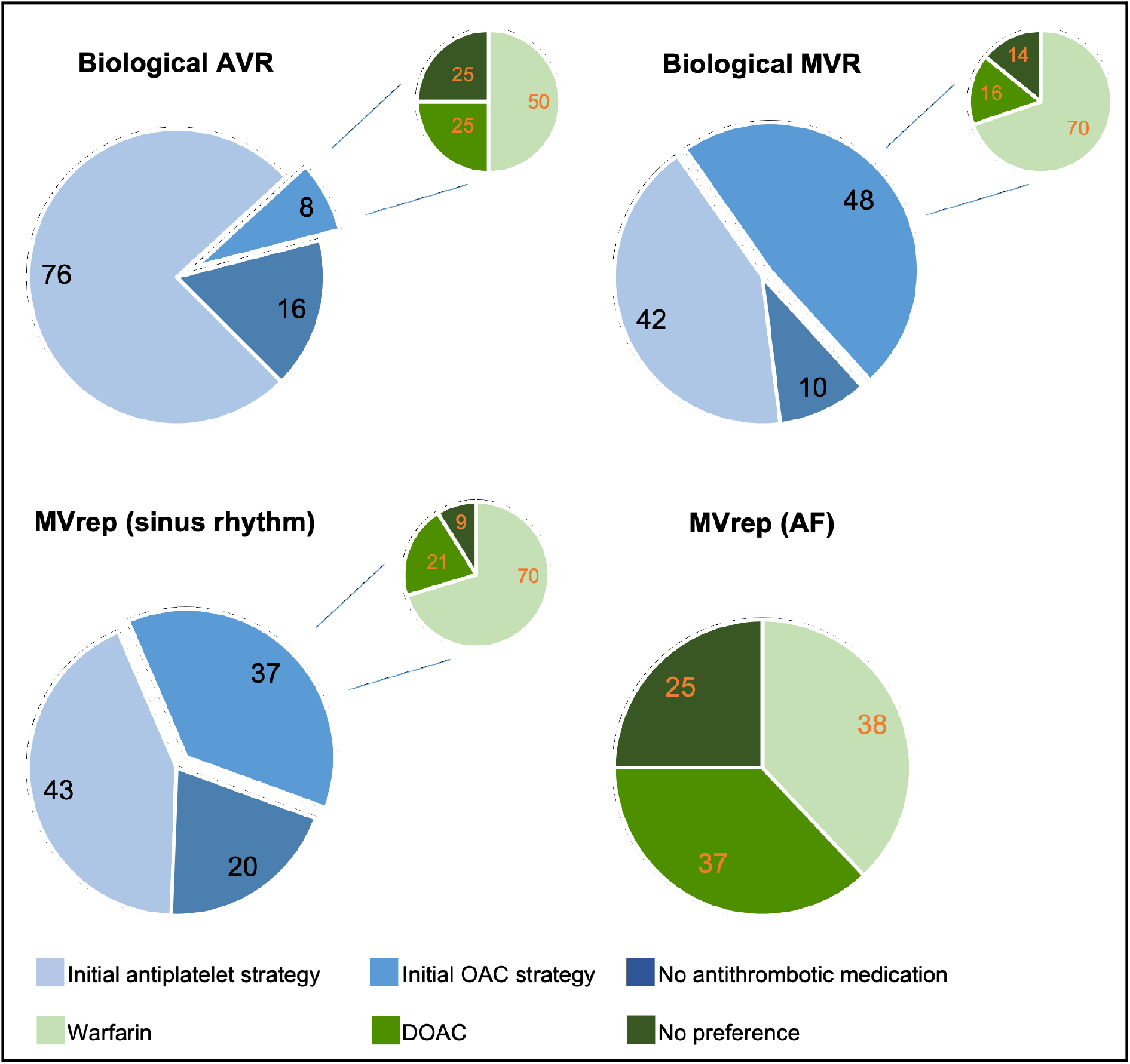
Initial antithrombotic strategy following bioprosthetic heart valve surgery or mitral valve repair. Number in slice denote percentage. AF, atrial fibrillation; AVR, aortic valve replacement; DOAC, direct oral anticoagulant; MVR, mitral valve replacement; MVrep, mitral valve repair; OAC, oral anticoagulant.

For patients undergoing bioprosthetic MVR, there was considerable variation in practice. There was similar use of an initial OAC (48%) or antiplatelet (42%) strategy, usually for 3 months. Two-thirds of surgeons opted to follow this initial therapy with lifelong antiplatelet therapy, whereas 23% of surgeons recommend some form of antithrombotic therapy (antiplatelet 10%, OAC 13%) for the first 3 months only, with no long-term treatment thereafter.

For patients undergoing mitral valve repair in sinus rhythm, 43% of surgeons recommended an initial antiplatelet strategy, with 37% opting for an initial OAC regime for 3 months. Only one-third of surgeons continued a lifelong antiplatelet agent thereafter. In patients undergoing MVrep with chronic AF, approximately equal proportions of surgeons prescribed warfarin (38%) or a direct oral anticoagulant (DOAC) (37%), with the remainder (25%) declaring no preference as to class of OAC used.

## DISCUSSION

Our cross-sectional survey of contemporary practice regarding use of antiplatelet and anticoagulant drugs after heart valve surgery in the UK has revealed clear inter-hospital and intra-hospital variation in practice. Some of the observed differences may be due to surgeons following different guidelines (ACC/AHA and ESC/EACTS guidelines differ in their recommendations) but there is also evidence of practice not supported by any guidelines.

We did not explore rationale for treatment strategies adopted, but posit that the lack of standard practice within departments may be due to individual surgical experience, comfort with a particular agent and lack of data to prompt a change in practice.

Thromboembolic complications leading to major cardiac or cerebrovascular events and death after bioprosthetic valve replacement or mitral valve repair are viewed as a significant risk by patients and clinicians, as are bleeding complications caused by anticoagulant and antiplatelet drugs used to prevent them. An absence of randomised trials, with discordant international guidelines and the emergence of DOACs, means optimal care – which could reduce unnecessary mortality and morbidity and improve quality of life after valve surgery – has remained elusive. In the recently conducted James Lind Alliance Priority setting exercise in adults undergoing cardiac surgery, patients consistently stated that the idea of such important decisions about post-operative therapy being based on clinician preference, and not on high quality research evidence, was alarming.^11^

The need to prevent thromboembolic complications must be balanced against the risk of major bleeding complications. For example, VKAs may be the most effective agents to reduce thromboembolic complications; however even when INR control is adequate, it is associated with a major bleeding complication rate of 1-3% per year.^12,13^ Bleeding complications can have a devastating impact on patients, including major morbidity and death. For example, the ORBIT-AF II study showed that major bleeding in patients on warfarin was associated with a 17% mortality and 47% hospitalisation rate.^14^ Similarly the ACTIVE-Warfarin and RE-LY trials indicated that the risk of death was increased 8-fold after an ischemic stroke, but 27-fold after a haemorrhagic stroke, and 5-fold after an extracranial bleeding event.^15,16^ In addition, trials such as ENGAGE-AF have demonstrate that reducing major bleeding reduces mortality.^17^

DOACS complicate the picture, with recent trials suggesting non-inferiority to warfarin following biological MVR but harm relative to warfarin in rheumatic mitral valve disease^13,18^ DOACs are sometimes preferred by patients and clinicians over warfarin because they do not require monitoring and have a lower risk of intracranial haemorrhage. However, they are more expensive, and these advantages may not be acceptable if they were not as effective as warfarin at also preventing thromboembolic complications. Dabigatran and apixaban have been shown to be ineffective and unsafe for patients with mechanical valve prostheses.^19-21^

Replacement valves in the mitral position have a higher risk of thromboembolism than in the aortic position. This may be due to a higher incidence of atrial fibrillation and left atrial enlargement and hence greater substrate for thrombosis. Therefore, early anticoagulation is thought to be beneficial with continuation of therapy based on risk factors (AF, dilated left atrium and left ventricular dysfunction).^10^

Our survey noted an increasing trend of using DOACs instead of VKAs as anticoagulation after biological valve replacements and mitral valve repair surgery. However, several recent publications have shown that DOACs and VKAs cannot be used interchangeably. The RIVER trial compared DOAC versus VKA in patients with AF undergoing bioprosthetic MVR, most of whom had rheumatic valve disease. This found that rivaroxaban was noninferior to warfarin with respect to the mean time until the primary outcome of death, major cardiovascular events, or major bleeding at 12 months.^13^ Conversely the INVICTUS trial has recently reported that among patients with rheumatic mitral valve disease and atrial fibrillation, VKA therapy led to a lower rate of a composite of cardiovascular events or death than rivaroxaban therapy, without a higher rate of bleeding compared to a DOAC.^18^

Subclinical valve thrombosis indicated by reduced leaflet mobility and hypo-attenuated leaflet thickening (HALT) has been identified in transcatheter and surgical aortic valve replacements on CT imaging. This has been reported to lead to cerebral embolic events.^3^ These abnormalities may resolve with anticoagulation and although we still do not know the clinical significance of HALT, this represents a potential indication for initial anticoagulation post-surgery. Defining the treatment for HALT after biological surgical valve replacement is an urgent priority.

In degenerative mitral disease, MVrep is preferred over replacement in international guidelines^9,10^ for lower morbidity, mortality, and improved durability. Despite a lower burden of prosthetic material, the incidence of early systemic thromboembolism remains high at 2.5% within 30 days and 4.2% at 6 months.^22^ The ESC/EACTS guidelines recommend three months of warfarin following mitral repair to allow endothelialisation of the annuloplasty ring. The evidence to support this recommendation, however, comes from a heterogenous population of mitral repairs and replacements.^10^ There are no recommendations in the ACC/AHA guidelines for antithrombotic management following MVrep, with the authors concluding there was not sufficient evidence to make a recommendation and highlighting the need for research.^9^

The recent National Institute for Health and Care Excellence (NICE) Guideline for Heart Valve Disease Presenting in Adults states for patient undergoing valve repair that ‘no evidence was identified comparing different anticoagulant and antiplatelet treatments in adults who have had valve repair. The committee made a recommendation for research comparing anticoagulant and antiplatelet treatments with placebo after valve repair’.^23^

Our survey revealed that for patients in sinus rhythm having MVrep, most surgeons in the UK favour antiplatelet therapy. For patients with AF having MVrep, VKA and DOACs were used with similar frequency.

## LIMITATIONS

There are certain strengths and limitations of this study that merit discussion. Strengths include the breadth of surgeons across the UK that responded, with answers received from 33 of the 36 surgical centres. However, we acknowledge that fewer than 50% of the contacted surgeons responded to the survey. Not all questions were completed by all respondents, since not all surgeons perform mitral valve repair surgery. The broad nature of questions precluded inclusion of specific details that may have impacted clinical decision-making for certain patients (e.g. left atrial size or whether left atrial appendage removal/occlusion was performed at the time of mitral valve repair).

## CONCLUSION

This contemporary cross-sectional survey has revealed great variation in clinical practice regarding use of antithrombotic drugs after heart valve surgery across the United Kingdom. Variation existed between different surgical centres but also between surgeons within the same centre, both with respect to agent(s) chosen and duration of therapy. These variations likely reflect the lack of robust data to guide decision-making and support the need for randomised studies to inform clinical practice.

## Data Availability

All data produced in the present study are available upon reasonable request to the authors

