## Supplemental material for "Antithrombotic Therapy After Heart Valve Surgery, Contemporary Practice in the United Kingdom"

Questions (1-10) refer to patients in sinus rhythm who do not have any other medical reason to take antiplatelet drugs (e.g. prior heart attack or stroke or intra-coronary stents) or anticoagulant drugs (e.g. atrial fibrillation, prior pulmonary emboli). Question 11 is a specific question about your choice of anticoagulant in patients with AF following mitral valve repair.

1. Is there a departmental policy in your hospital on the use of anti-thrombotic drugs after bioprosthesis implantation?
   - Yes
   - No
2. Do you prescribe/recommend anti-thrombotic drugs after bioprosthetic aortic valve replacement?
   - Yes
   - No
3. If Yes
   - 3 months antiplatelet therapy, then stop
   - 3 months anticoagulation, then stop
   - 3 months anticoagulation, then lifelong antiplatelet drug
   - Lifelong antiplatelet drug
   - Other:
4. If an anticoagulant is used is this

- NOAC (i.e. dabigatran / rivaroxaban / apixaban / edoxaban)
- Warfarin (or other Vit K antagonist)
- Either (no preference)

5. Do you prescribe/recommend anti-thrombotic drugs after bioprosthetic mitral valve replacement?

- Yes
- No

6. If Yes

- 3 months antiplatelet therapy, then stop
- 3 months anticoagulation, then stop
- 3 months anticoagulation, then lifelong antiplatelet drug
- Lifelong antiplatelet drug
- Other:

7. If an anticoagulant is used is this

- NOAC (i.e. dabigatran / rivaroxaban / apixaban / edoxaban)
- Warfarin (or other Vit K antagonist)
- Either (no preference)

8. Do you prescribe/recommend anti-thrombotic drugs after mitral valve repair?

- Yes
- No

9. If Yes

- 3 months anticoagulation, then stop
- 3 months anticoagulation, then lifelong antiplatelet drug
- Lifelong antiplatelet drug
- Other:

10. If an anticoagulant is used is this

- NOAC (i.e. dabigatran / rivaroxaban / apixaban / edoxaban)
- Warfarin (or other Vit K antagonist)
- Either (no preference)

11. For patients who have mitral valve repair and remain in chronic atrial fibrillation, what form of anticoagulation do you recommend?

- Warfarin (or other Vit K antagonist)
- NOAC (i.e. dabigatran / rivaroxaban / apixaban / edoxaban)
- Either (no preference)
